# A Phenome Wide Association Study of Severe COVID-19 Genetic Risk Variants

**DOI:** 10.1101/2021.12.08.21267433

**Authors:** Jessica A. Regan, Jawan Abdulrahim, Nathan Bihlmeyer, Carol Haynes, Lydia Coulter Kwee, Manesh Patel, Svati H. Shah

## Abstract

**Background:** Genetic loci associated with risk of severe COVID-19 infection have been identified and individuals with complicated COVID-19 infections often have multiple comorbidities.

**Objective:** Identify known and unidentified comorbidities associated with genetic loci linked to risk of severe COVID-19 infection.

**Methods:** A Phenome Wide Association Study (PheWAS) was conducted in 247,448 unrelated, white individuals from the UK Biobank to test the association of 1,402 unique phenotypes with ten genome-wide significant severe-COVID risk single nucleotide polymorphisms (SNP) identified from prior studies. A validation PheWAS was conducted in 2,247 white individuals from the CATHGEN.

**Results:** Four of the ten tested genetic loci showed significant phenotypic associations in UK Biobank after FDR adjustment. Vascular dementia significantly associated with rs7271165 near *TMEM65* on 8q24.13 in individuals with the C risk allele (OR 5.66 [95% CI 2.21-11.85], q=0.049). We identified 40 novel phenotype associations with rs657152 on 9q34.2 coinciding with the *ABO* gene with individuals with the A COVID risk allele having higher odds of heart failure (OR 1.09 [95% CI 1.03-1.14], q=0.004), diabetes mellitus (OR 1.05 [95% CI 1.02-1.07], q=0.004) and hypercholesterolemia (OR 1.04 [95% CI 1.02-1.06], q=6.3×10^−5^). Eight phenotypes associated with rs1819040 near *KANSL1* on 17q21.31 in individuals with the A risk allele including atrial fibrillation and flutter (OR 1.07 [95% CI 1.04-1.10], q=0.0084) and pulmonary fibrosis (OR 0.80 [95% CI 0.71-0.89], q=0.035). Ten novel phenotypic associations were identified in association with rs74956615 on 19p13.2 near the *TYK2* gene including individuals with the A COVID risk allele having lower odds of psoriatic arthropathy (OR 0.31 [95% CI 0.20-0.47], q=4.5×10^−5^), rheumatoid arthritis (OR 0.83 [95% CI 0.64-0.83], p=1.4×10^−6^) and thyrotoxicosis with or without goiter (OR 0.77 [95% CI 0.68-0.87], p-6.9×10^−5^). Two associations for rs1819040 (*KANSL1*) and seven associations for rs74956615 (*TYK2*) validated in CATHGEN.

**Conclusions:** Using a broad PheWAS approach in a large discovery and validation cohort, we have identified novel phenotypic associations with risk alleles for severe COVID-19 infection. Interestingly, the ABO locus was associated with comorbidities that are also risk factors for severe COVID-19 infection, suggesting that this locus has pleiotropic effects and provides a potential mechanism for this association. The 19p13 locus was associated with lower risk of autoimmune disease, these findings may have broad implications for the importance of multiple comorbidities across both infectious and non-infectious diseases and may provide insight in the molecular function of the genes near these genetic risk loci.

## Introduction

There is marked heterogeneity in the clinical manifestations of coronavirus disease 2019 (COVID-19), which is caused by infection with the severe acute respiratory syndrome coronavirus 2 (SARS-CoV-2). Symptoms can range from mild-flu like symptoms to severe respiratory failure requiring supplemental oxygen, intubation or intensive care unit (ICU) care and multiple distinct cardiovascular complications have also been identified^1^. Demographic factors and existing clinical comorbidities are associated with severe COVID-19. For example, age, male gender, Black and South Asian ancestry, diabetes, obesity and chronic lung disease are associated with increased risk of COVID-19-related mortality^2^.

The biologic underpinnings for the heterogeneity in increased risk with these clinical comorbidities is unclear, however, inherited genetic factors have been associated with severe COVID-19. In June 2020, using a genomewide association study (GWAS) analyses of common genetic variants, the Severe COVID GWAS Group first identified two single nucleotide polymorphisms (SNPs) with genome-wide significance for severe COVID infection using a meta-analysis of 1,610 participants with severe COVID-19 with respiratory failure and 2,205 healthy controls across seven hospitals in Italy and Spain^3^. The first SNP was rs11385942 at locus 3p21.31 with a signal spanning multiple genes including chemokine receptors: *SLC6A20, LZTFL1, CCR9, FYCO1, CXRC6* and *XCR1*. The second SNP was rs657152 at locus 9q34.2, coinciding with the *ABO* blood locus group, with analyses showing greater risk of severe COVID in type A blood carriers and protective effects in individuals with type O blood group. Further, a small study of 4 young male patients without chronic disease treated for severe COVID-19 requiring mechanical ventilation and ICU care analyzed rare variants and found loss-of-function variants in *TLR7* on the X-chromosome with associated impairment in type I and type II interferon (IFN) responses^4^. In December 2020, the Genetics Of Mortality In Critical Care (GenOMICC) genome-wide association study of 2,244 critically ill patients across 208 United Kingdom (UK) ICUs identified and replicated four additional genome-wide significant signals^5^. Most recently in July, 2021, the COVID-19 Host Genetics Initiative has identified ten distinct genome-wide significant loci associated with severe COVID-19 and confirmed the findings of these earlier studies^6^.

Given that greater comorbidities have also been observed in patients with severe COVID-19 infection we aimed to identify association between a wide range of comorbidities for these same genetic loci associated with severe COVID-19, with the goal of better understanding potential genetic risk of severe COVID-19 mediated by these variants. Phenome-wide association study (PheWAS) has emerged as an unbiased approach to identify novel associations of previously identified, disease-associated genetic variants, across many phenotypes. One such PheWAS study has been conducted for the 3p21.31 locus^7^, however, additional phenotypic associations and broader implications of risk for additional identified COVID-19 genetic loci have not yet been described.

## Methods

### Study Populations

The UK Biobank is a prospective population-based cohort with deep genetic and phenotypic data collected on 500,000 individuals 40-69 years old at recruitment (2006-2010) across the United Kingdom^8^. For this study, we selected a discovery cohort composed of 247,488 unrelated white British ancestry UK Biobank participants with both high-quality genotype and phenotype data available. Genetic data were obtained from either the UK Biobank Axiom array from Affymetrix or UK BiLEVE Axiom array and are reported in Genome Reference Consortium Human Reference 37 (GRCh37). Imputation was performed by UK Biobank using a merged panel of the Haplotype Reference Consortium (HRC) panel, the UK10K panel and the 1000 Genome Phase 3 panel. Phenotype data was derived from International Classification of Diseases (ICD) codes from primary care data, hospitalizations and death-related data in the UK Biobank. Data for this project was approved and accessed through approved UK Biobank applications (Application ID: 48785 and 65043).

The validation cohort consisted of individuals from the CATHGEN study, a study of 9334 sequential individuals who underwent cardiac catherization at Duke University Medical Center (Durham, NC) between 2001-2010^9^. The validation cohort was comprised of 2,247 self-reported white individuals from the CATHGEN study. Genotype data were obtained using the Illumina Human Omni1-Quad Infinium Bead Chip, and imputed with Minimac4 using 1000G phase 3 reference panels and are reported in GRCh37^10^. Phenotype data was derived from electronic health record data from 2001-2020.

Both studies have human subjects research approval from the Duke Institutional Review Board (IRB) and all participants provided informed consent.

### Selection of COVID-19 Risk Genetic Variants

Ten single nucleotide polymorphisms (SNP) were selected for use from prior studies of COVID-19. Specifically, amongst the other fine-mapped SNPs for each loci, consideration was given to linkage disequilibrium, availability of the variant data in each cohort and minor allele frequency (MAF) for inclusion, with the goal of selecting only one SNP from each locus. The ten severe COVID-19 associated loci studied here are: rs11385942 (3p21.31 composed of a six gene cluster, risk allele GA, MAF=0.07), rs1886814 (6p21.1, closest gene *FOXP4*, risk allele C, MAF=0.03), rs72711165 (8q24.13, closest gene *TMEM65*, risk allele C, MAF=0.01), rs657152 (9q34.2, closest gene *ABO*, risk allele T, MAF=0.35),rs10735079 (12q24.13, closest gene *OAS1*, risk allele A, MAF=0.36), rs1819040 (17q21.31, closest gene *KANSL1*, risk allele A, MAF=0.22), rs77534576 (17q21.33, closest gene *TAC4*, risk allele T, MAF=0.03), rs74956616 (19p13.2, closest gene *TYK2*, risk allele A, MAF=0.05), rs210969 (19p13.3, closest gene *DPP9*, risk allele A, MAF=0.32) and rs2236757 (21q22.2, closest gene *IFNAR2*, risk allele C, MAF=0.28). rs11385942 and rs657152 were the lead SNPs for the two loci from the initial Italian and Spanish GWAS^3^. The four SNPs initially identified and replicated by GenOMICC were rs10735079, rs74956616 (19p13.2, *TYK2*, MAF=0.05), rs210969 (19p13.3, *DPP9*, MAF=0.32) and rs2236757 (21q22.2, *IFNAR2*, MAF=0.28)^5^. The remaining four SNPs were most recently identified through the COVID-19 Host Genetics Initiative.

### Statistical Analysis

Outcomes were defined by the PheCODE scheme using ICD 9 and ICD 10 codes to identify phenotypes, where incident and prevalent cases were included for both studies^11, 12^. The R PheWAS package was used for all analyses^13^. All outcomes with <20 cases were excluded resulting in 1,403 phenotypes assessed in UK Biobank and 1,002 phenotypes in CATHGEN. The full list of phenotypes can be found in Supplementary Table 1. Logistic regression was performed for disease outcomes and all analyses were adjusted for age, genotyping array, gender and the first five genetic principal components. In the UK Biobank discovery cohort, significant associations were considered at an FDR adjusted p-value (q-value) and nominal associations were considered at p<0.05. In the CATHGEN cohort, nominal association at p<0.1 was considered for validation. All analyses were performed in using R v4.0.2.

## Results

### Novel phenotypic associations with four severe COVID-19 risk alleles

Four out of the ten tested severe COVID-19-associated genetic risk loci showed significant phenotypic associations in the UK Biobank dataset after FDR adjustment. First, vascular dementia significantly associated with rs72711165 (*TMEM65*) (OR 5.66 [95% CI 2.21-11.85], q=0.049, but did not validate in CATHGEN (**Figure 1A, Supplemental Table 3**). We identified 40 significant phenotype associations for rs657152 after FDR adjustment, 26 of which have not been previously described^11^. Notable associations include greater odds of heart failure (OR 1.09 [95% CI 1.03-1.14], q=0.046), diabetes mellitus (OR 1.05 [95% CI 1.02-1.07], q=0.004 and Hypercholesterolemia (OR 1.04 [95% CI 1.02-1.06], p=q=0.004) and lower odds of gastrointestinal disorders including duodenal ulcer (OR 0.88 [95% CI 0.84-0.92], q=6.3×10^−5^, **Figure 1B, Supplemental Table 4**) with non-group O blood types. However, none of these novel findings validated in the CATHGEN cohort.

**Figure 1.**
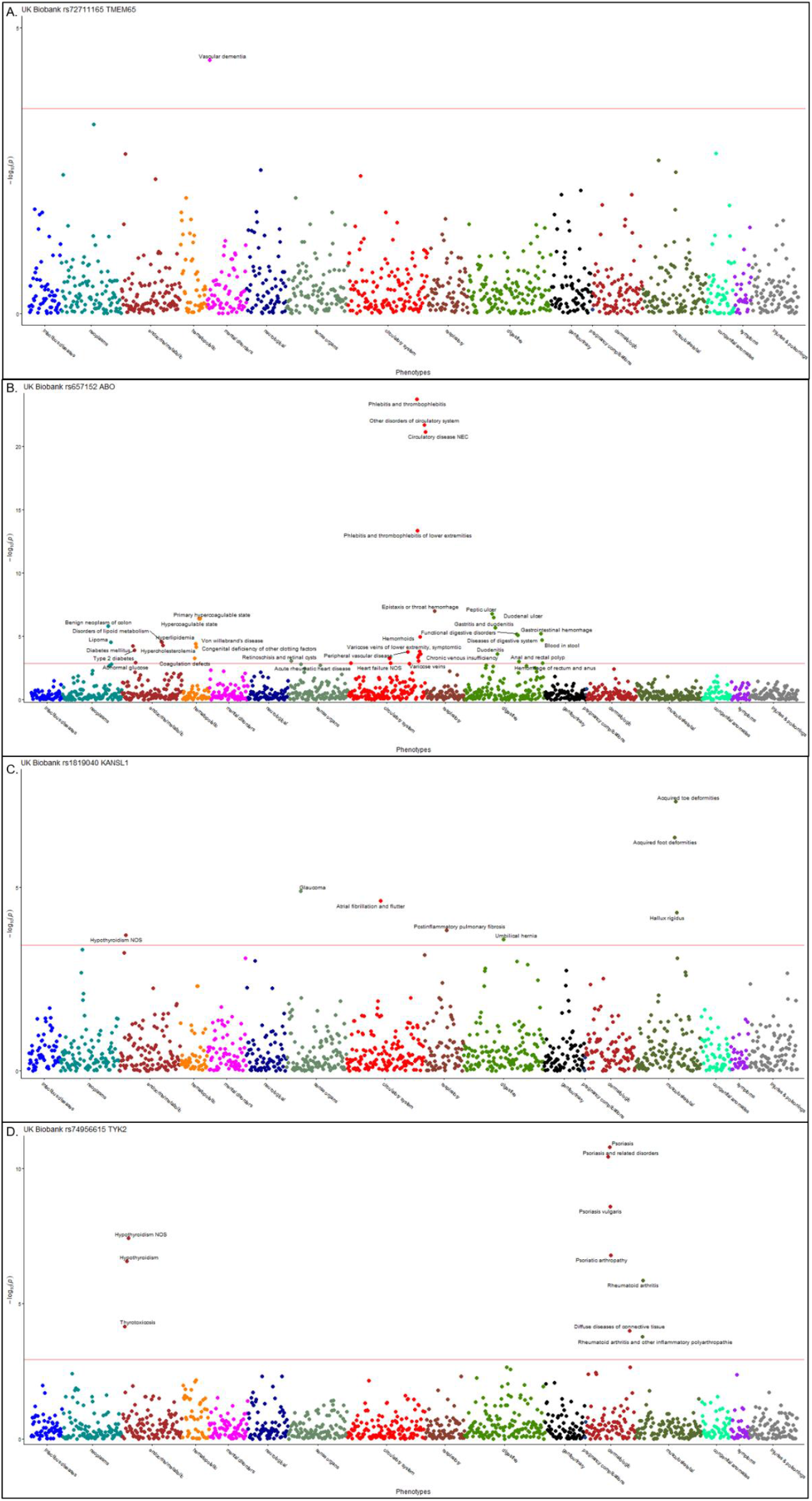
Significant Phenotypic Associations with COVID-19 risk alleles in UK Biobank. Shown are the results of the significant findings of the phenome-wide association study for severe COVID-19 single nucleotide polymorphisms. The x-axes correspond to the different groups of phenotypes analyzed and the y-axes correspond to the negative logarithm p-values for these analyses. The red line corresponds to statistical significance level at a false discovery rate<0.05, phenotypes meeting statistical significance are annotated. **A**. Results for rs72711165 *TMEM65* **B**. Results for rs657152 *ABO*. **C**. Results for rs1819040 *KANSL1* **D**. Results for rs74956615 *TYK2*.

Eight phenotypes associated with rs1819040 (*KANSL1*), including atrial fibrillation and flutter (OR 1.07 [95% CI 1.04-1.10], q=0.0084) and pulmonary fibrosis (OR 0.80 [95% CI 0.71-0.89], q=0.035) (**Figure 1C, Supplemental Table 6**). Only glaucoma validated in CATHGEN (p<0.1). Ten phenotypes were significantly associated with rs74956615 (*TYK2*) in UK Biobank after FDR adjustment, all with lower odds associated with the COVID-19 risk allele (**Figure 1D, Supplemental Table 8**), including psoriatic arthropathy (OR 0.31 [95% CI 0.20-0.47], q=4.5×10^−5^), rheumatoid arthritis (OR 0.83 [95% CI 0.64-0.83], q=0.0003) and thyrotoxicosis (OR 0.77 [95% CI 0.68-0.87], q=0.01). Seven phenotypes nominally validated in CATHGEN, including psoriasis, rheumatoid arthritis and hypothyroidism (p<0.1).

### Nominal associations with other severe-COVID-19 risk alleles

No phenotypes were associated with rs11385942 at the 3p21.31 locus at FDR level adjustment; however, 68 out 1,403 of phenotypes were nominally associated in the UK Biobank cohort (p<0.05, **Supplemental Table 1**). These are all novel findings, as no phenotypic associations have ever been reported before for rs11385942 beyond severe COVID-19. Some of the most significant phenotypes included obstructive chronic bronchitis (OR of disease associated with carriage of one COVID-19 risk allele was 1.18 [95% CI 1.07-1.29], p=0.0006), hypertensive heart disease (OR 1.50 [95% CI 1.24-2.31], p=0.0007) and atrial fibrillation (OR 1.13 [95% CI 1.00-1.26], p=0.04). Carriers of the COVID-19 risk allele for rs11385942 had lower odds of Eosinophilia (OR 0.43 [95% CI 0.18-0.85], p=0.03) and Elevated sedimentation rate (OR 0.60 [95% CI 0.40-0.96], p=0.0008). One of these 68 associations for memory loss validated in the CATHGEN cohort (p<0.05) and one other for Neurologic disorders, nominally validated (p<0.1).

No phenotypes were associated with rs1886814 at the 6p21.1 loci (*FOXP4*) after FDR adjustment. 63 phenotypes were nominally associated with this COVID-19 risk allele, including greater odds of nephrotic syndrome (OR 1.54 [95% CI 1.14-2.03], p=0.004) and chronic lymphocytic thyroiditis (OR 1.73 [95% CI 1.08-2.62], p=0.01), as well as lower odds of empyema (OR 0.69 [95% CI 0.49-0.94], p=0.02) and viral pneumonia (OR 0.12 [95% CI 0.01-0.58], p=0.049, **Supplemental Table 2**). None of these findings validated in CATHGEN.

No phenotypes met FDR-threshold for significance with rs10735079 (*OAS1*) 85 phenotypes were nominally associated, where individuals with the COVID-19 risk allele had a lower odds of circulatory disorders including, polyarteritis nodosa (OR 0.64 [95% CI 0.44-0.95], p=0.02), peripheral vascular disease (OR 0.96 [95% CI 0.92-1.00, p=0.04), and ischemic heart disease (OR 0.97 [95% CI 0.95-0.99], p=0.01), as well as lower odds of psoriasis and related disorders (OR 0.94 [95% CI 0.01-0.98], p=0.002), hypothyroidism (OR 0.96 [95% CI 0.94-0.98], p=0.002] and renal failure (OR 0.97 [95% CI 0.94-1.00], p=0.02, **Supplemental Table 5**). rs10735079 was associated with greater odds of tuberculosis (OR 1.37 [95% CI 1.03-1.85], p=0.03) and other disease of lung (OR 1.13 [95% 1.02-1.25], p=0.02). One phenotypic association for rs10735079 nominally validated in the CATHGEN cohort for lower odds of benign neoplasm of colon (p<0.1).

No phenotypes were significantly associated with rs77534576 (*TAC4*) after FDR adjustment. 76 phenotypes were nominally associated, with individuals carrying the COVID-19 risk allele having a greater odds of acute renal failure (OR 1.23 [95% CI 1.10-1.37], p=0.0003] and Hodgkin’s disease (OR 1.84 [95% CI 1.20-2.70], p=0.003), as well as both hypertensive complications (OR 2.27 [95% CI 1.39-3.74], p=0.0004) and hypotension (OR 1.20 [95 % CI 1.09-1.32], p=0.0003, **Supplemental Table 7**). Two phenotypes, including postinflammatory pulmonary fibrosis (OR 1.32 [95% CI 1.01-1.68], p=0.03) validated in CATHGEN (p<0.1).

In UK Biobank, there were no FDR significant phenotypic associations with rs2109069 *(DPP9*). 72 phenotypes were nominally associated including other specified diffuse diseases of connective tissue (OR 2.05 [95% CI 1.44-2.92], p=6.4×10^−5^), rheumatoid arthritis (OR 1.07 [95% CI 1.01-1.13], p=0.02), diabetes mellitus (OR 1.03 [95% CI 1.00-1.05], p=0.02), Respiratory failure (OR 1.07 [95% CI 1.01-1.13], p=0.03, **Supplemental Table 9**). One phenotype validated in CATHGEN for lower odds of Immunity deficiency associated with the COVID-19 risk allele (p<0.05).

No phenotypes were significantly associated with rs2236757 in UK Biobank (**Supplemental Table 10**). 83 phenotypes were nominally associated including, greater odds of viral pneumonia (OR 1.61 [95% CI 1.25-2.12, p=0.0004) and thyroiditis (OR 1.20 [95% CI 1.04=1.38], p=0.01). However, individuals carrying the COVID-19 risk allele had lower odds of bacterial infection (OR 0.96 [95% CI 0.92-0.99], p=0.01), pneumonia (OR 0.97 [95% CI 0.94-1.00], p=0.04) and viral infection (OR 0.96 [95% CI 0.94-1.00], p=0.03). rs2236757 was also associated with lower odds of respiratory abnormalities (OR 0.90 [95% CI 0.81-0.99], p=0.04), hypertensive heart and/or renal disease (OR 0.91 [95% CI 0.84-0.99], p=0.02) and polymyalgia rheumatic (OR 0.91 [95% CI 0.85-0.98], p=0.01). Five phenotypes validated in CATHGEN including a lower odds of bacterial infection (p<0.05).

## Conclusions

Using an unbiased PheWAS approach to clinical diagnoses in a large dataset of genetic and electronic health record data, we have identified novel phenotypic associations with the risk alleles from four of ten loci previously identified as associated with severe COVID-19 infection. These associations could suggest that individuals carrying these genetic markers, known for their role in blood traits, host anti-viral response and inflammation, may have modified risk of cardiovascular disease, as well as auto-immune and inflammatory disorders including arthropathies and endocrinopathies, which in turn increases risk of severe COVID-19. Alternatively, these genetic risk loci may have pleiotropic effects on these diseases and on COVID-19 related complications.

No prior phenotypic associations are published for the rs72711165 SNP near *TMEM65. TMEM65* is a mitochondrial inner-membrane protein that may play a role in mitochondrial respiration and cardiac development and function. Mutations in *TMEM65* have been described to cause mitochondrial myopathy and neurologic disease^14^. Direct mechanisms related to the association with vascular dementia identified here are unclear, but warrant further investigation.

Prior to being identified as a risk variant for severe COVID-19, rs657152 within the *ABO* blood locus group, had been associated with hypercoagulable state, arterial embolism and thrombosis and other disorders of circulatory system^11^. We validated these previously reported associations for the rs657152 SNP and identified novel associations including with greater odds of heart failure, diabetes mellitus, and hypercholesterolemia and lower odds of gastrointestinal disorders including duodenal ulcer and duodenitis. Genetic predisposition for these cardiovascular and endocrine phenotypes may amplify the risk of adverse COVID-19 outcomes but may also have broader long-term health implications^15^. Taken together these associations add support to risk factors contributing to a hypercoagulable state, as both the rs657152 risk allele and COVID-19 infection itself may increase risk of via multiple mechanisms of thrombosis^16^.

Mutations in *KANSL1* are known to cause neurodevelopmental delay disorders described within 17q21.31 deletion syndrome or Koolen-de Vries syndrome^17^. *KANSL1* plays a role in histone acetylation, microtubule stabilization and mitochondrial respiration^18^. Here, we identified novel associations with the rs1819040 SNP near KANSL1, including greater odds of atrial fibrillation, hypothyroidism and glaucoma, and interestingly, lower odds of postinflammatory pulmonary fibrosis. Biologic mechanisms linking these associations, as well as the risk of severe COVID-19, warrant further study.

We also found that rs74956615 near the *TYK2* gene was associated with lower odds of psoriasis and related disorders, rheumatoid arthritis and thyrotoxicosis, as well as greater odds of tobacco use disorder. Adding strength to the results for rs74956615, these findings nominally validated in the CATHGEN cohort. *TYK2*, a member of the Janus Kinase (JAK) family, is involved in interleukin-23 (IL-23) signaling, a cascade associated with psoriasis via Th17 responses and IFN-α signaling. Therapeutic targeting of JAK signaling and *TYK2* is implicated in auto-immune and inflammatory diseases including both psoriasis and rheumatoid arthritis^19^. Nine prior SNPs in *TYK2* have been reported in association with autoimmune diseases including psoriasis, rheumatoid arthritis, systemic lupus erythematosus (SLE) and inflammatory bowel disease (IBD), however, these studies have had mixed directions of effect. A recent systematic review and meta-analysis identified protective effects against autoimmune disease for five *TYK2* SNPs and risk for SLE associated with one^20^. Rare coding variants found to have protective effects have been associated with reductions in IL-23 and IFN-α signaling associated with these rare coding variants^21^. Here for first time we show decreased odds of psoriasis associated with rs74956615, which may implicate a distinct impact of this allele on *TYK2* gene function from what has been previously identified in prior GWAS analysis of psoriasis. Notably previous investigators studying the protective impact of *TYK2* variants on autoimmune disease did not identify pleiotropic effects via PheWAS analyses^22^ and the associations of *TYK2* and thyroid disease found in the present analyses have not been previously reported, however the utilization of the UK Biobank cohort represents the largest analysis of *TYK2* variants to date. Our study design does not allow for more detailed confirmation of whether the reported cases of hypothyroidism may have been autoimmune in etiology.

Though findings did not reach prespecified significance thresholds in the present analyses, the other identified COVID-19-related genetic variants suggest the importance of host antiviral defense mechanisms and inflammatory signaling. Zhou et al performed a PheWAS of 310,999 European individuals in the UK Biobank and identified blood cell traits including monocyte and eosinophil count to be associated with the 3p21.31 locus^7^. Though findings at other loci were only nominally associated, these findings may still be suggestive of relevant phenotypic and molecular pathways for these genetic loci and warrant further investigation in more clinical and pre-clinical models. Nominal associations for lower odds of eosinophilia corroborate the recent findings by Zhou et al for the 3p21.31 SNP rs11385942^7^.

Limitations to this study include the limited sample size in the validation cohort which may make us under powered to fully validate results. Additionally, these analyses were only conducted in individuals of European ancestry based on the ancestry breakdown in the cohorts available. Clinical research should continue to focus efforts to recruit diverse patient populations to help us generate research findings broadly applicable to patients, particularly in the context of health disparities observed in COVID-19.

## Supporting information

STROBE Checklist

## Data Availability

All data produced in the present work are contained in the manuscript and supplement

## Acknowledgements

We would like to thank the UK Biobank and CATHGEN participants whose data contributed to this work.

## Abbreviations

COVID-19: SARS-coronaravirus-19
UK Biobank: United Kingdom Biobank
PheWAS: Phenome Wide Association Study
CATHGEN: Catheterization Genetics
FDR: False discovery rate

## Notes

**Funding** J.A.R. is supported by a grant from the National Heart, Lung and Blood Institute (1R38HL143612).

### Competing Interest Statement

The authors have declared no competing interest.

### Funding Statement

Dr. Regan received funding from 1R38HL143612.

### Author Declarations

Ethics commmittee/IRB of Duke University gave ethical approval for this work

